# Projected burden of alcohol-associated liver disease in China, 2020-2050: A microsimulation modeling study

**DOI:** 10.64898/2026.08.19.26360748

**Authors:** Qingfeng Niu, Mingzhu Su, Leshi Liang, Zhaoyi Che, Qiang Zhu, Fei Wang, Jia Xiao

## Abstract

**Background:** Alcohol-associated liver disease (ALD) has emerged as a major cause of chronic liver disease and liver-related mortality in China. This study aimed to project the future burden of ALD in Chinese adults from 2020 to 2050, including prevalence of ALD, number of alcoholic steatohepatitis (ASH) cases, incident hepatocellular carcinoma (HCC) cases, liver transplantation (LT) demand, liver-related deaths, and disability-adjusted life years (DALYs).

**Methods:** We developed an agent-based state-transition microsimulation model with yearly cycles and a lifetime horizon. The model simulated 5,678,912 representative Chinese adults (mean age 36.2 years, 51.2% male). Health states included no steatosis, alcohol-associated steatotic liver, ASH, fibrosis stages F0-F4, decompensated cirrhosis, HCC, LT, and liver-related death. Model inputs were derived from the China Kadoorie Biobank, Global Burden of Disease Study 2021, China’s national surveys, published meta-analyses, and transplant registry data. Projections incorporated demographic shifts, alcohol consumption trends, and calibrated transition probabilities. Uncertainty was assessed via 1,000 Monte Carlo simulations generating 95% uncertainty intervals.

**Results:** ALD prevalence was projected to increase from 4.8% (55 million individuals) in 2020 to 8.5% (94 million individuals) by 2050. ASH cases rose from approximately 18 million to 20 million. Annual incident HCC cases nearly doubled from 20,500 in 2020-2025 to 45,200 by 2046-2050. LT demand quadrupled from 2,300 to 9,800 cases. Liver-related deaths increased from 50,000 in 2020 to 85,000 in 2050, while DALYs rose from 1.5 million to 2.6 million.

**Conclusions:** In the absence of strengthened alcohol control policies, ALD will impose a substantial and growing burden on China’s health system by 2050, with marked increases in HCC incidence, LT demand, and liver-related mortality.

## Background

The alcohol-associated liver disease (ALD), previously termed alcoholic liver disease, has become a growing global public health challenge, driven by rising alcohol consumption in many regions ^[1, 2]^. In China, *per capita* alcohol consumption increased from 4.1 liters of pure alcohol in 2005 to 7.2 liters in 2016, positioning ALD alongside metabolic dysfunction-associated steatotic liver disease (MASLD) as a major contributor to the national chronic liver disease burden ^[3, 4]^. By 2021, an estimated 6.4 million Chinese individuals were living with alcohol-related cirrhosis, and alcohol-attributable hepatocellular carcinoma (HCC) contributing approximately 20,500 new cases annually ^[5, 6]^. Individuals with ALD have elevated risks of alcoholic steatohepatitis (ASH), end-stage liver diseases, and liver transplantation (LT). With improved control of viral hepatitis, ALD is projected to become a leading indication for LT ^[7, 8]^. This study uses the updated term ALD to better reflect its metabolic and inflammatory components, while approximately 99% of cases previously classified as alcoholic liver disease align with current definitions ^[9]^.

Against this backdrop of escalating disease burden, heavy episodic drinking (binge drinking) and sustained heavy consumption accelerate the progression of ALD, with synergistic risks amplified by highly prevalent comorbidities including diabetes and obesity in China’s aging population ^[10]^. Although lifestyle interventions, including reduced alcohol intake, increased exercise, and emerging pharmacotherapies can improve clinical outcomes, access to these modifications remains limited nationwide ^[11]^. Accurate quantification of the clinical burden of ALD, especially the population eligible for advanced care (i.e., ASH with fibrosis stage ≥F2), is essential for health system preparedness and policy planning. Yet existing estimates vary considerably, hindering effective public health strategy. Prior studies using Markov cohort models have projected a rising burden of ALD prevalence and related outcomes in China and other settings, but these approaches typically rely on aggregate transition probabilities and homogeneous cohorts, limiting their ability to fully account for critical population heterogeneity, such as individual-level variations in age, sex, drinking patterns, comorbidities, and dynamic demographic shifts ^[5, 12]^. Using a microsimulation framework integrated with the latest evidence on disease progression, alcohol use patterns, and demographic transitions, this study aimed to project the comprehensive burden of ALD among Chinese population from 2020 to 2050, including the prevalence of ASH, HCC, LT, and liver-related mortality.

## Methods

### Study design

This decision analytical modeling study was reported following the Strengthening the Reporting of Empirical Simulation Studies (STRESS) guidelines ^[13]^. We developed an agent-based state transition model with a yearly cycle and lifetime time horizon. This approach was used to capture heterogeneity within the population, account for individual-level variation, and track the impact of that variation on individual outcomes, leading to a more accurate projection within a population ^[14]^. The model has two integrated components.

The first component established a synthetic population representative of the age and sex distribution of the Chinese population in 2020, with annual simulation through 2050. At baseline, we simulated 5,678,912 individuals (mean age 36.2 years; 51.2% male). This sample represented approximately 0.4% of the total Chinese adult population in 2020 (approximately 1.42 billion) extracted from medium variant of UN World Population Prospects (UNWPP) 2024 and was determined via internal pilot testing to satisfy two criteria: (1) adequate statistical precision to produce narrow 95% uncertainty intervals (UI) for low-probability events including HCC incidence (1%-4% per year) and liver transplantation (0.1%-0.5% per year); and (2) computational tractability for 1,000 Monte Carlo simulations. The precise cohort size was a practical, rounded value obtained by downscaling the full national adult population while preserving the age-sex structure specified in UNWPP 2024 and Chinese national statistics. Demographic dynamics-including births, migration, and mortality-were updated annually using UNWPP 2024 and Chinese national statistics.

The second component simulated the natural history of ALD among adults aged ≥18 years. Mutually exclusive health states included (Figure 1): no steatosis, alcohol-associated steatotic liver (ASL; simple steatosis), ASH, fibrosis (with or without ASH), compensated cirrhosis, decompensated cirrhosis (DC), HCC, LT, and liver-related death. Fibrosis was staged from F0 (no fibrosis) to F4 (cirrhosis). Each individual occupied exactly one health state at any time. At each annual cycle, individuals could either remain in their current state or transition to another state according to prespecified probabilities. ASL and ASH were modeled as parallel disease states, with bidirectional transitions permitted between steatosis and ASH at fibrosis stages F0-F2. Liver-related mortality encompassed deaths attributable to HCC, DC, and post-LT complications.

**Figure 1.**
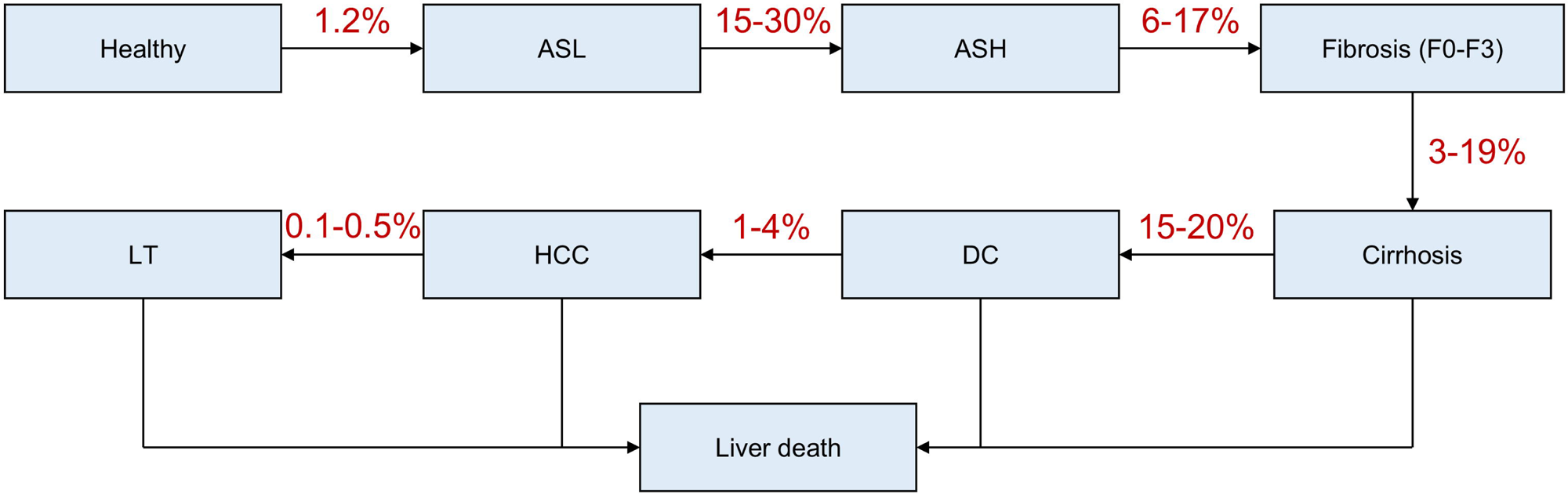
Schematic diagram of the agent-based state-transition model for alcohol-associated liver disease (ALD) in Chinese adults. Health states include no steatosis, alcohol-associated steatotic liver (ASL), alcoholic steatohepatitis (ASH), fibrosis stages F0–F3, cirrhosis, decompensated cirrhosis (DC), hepatocellular carcinoma (HCC), liver transplantation (LT), and death. Key annual transition probabilities are labeled on the arrows.

### Model inputs, data sources, and assumptions

All model inputs were derived from high-quality, peer-reviewed sources. The specific contributions of each data source are detailed in Table S1 ^[15–25]^. Detailed model inputs include: (1) age- and sex-specific alcohol consumption patterns, prevalence of hazardous drinking, and genetic evidence of causality (HRs for cirrhosis and gout) were extracted from China Kadoorie Biobank data; (2) baseline alcohol-attributable cirrhosis deaths, DALYs, and mortality rates for calibration were supplied by Global Burden of Disease (GBD) 2021 data; (3) drinking prevalence, urban-rural disparities, and low female drinking rates of Chinese adults were informed by CHARLS 2015-2020 waves; (4) histological progression rates of each stage of ALD and ASH proportions in biopsy cohorts were extracted from published meta-analyses; (5) LT probabilities, population projections, and life tables for DALY calculation were from Chinese national transplant registry and UNWPP 2024. For unknown or uncertain parameters, we used calibration against observed targets (e.g., 2020 prevalence from national surveys and GBD 2021 mortality). All assumptions are listed in Table S1.

### Mathematical modeling framework

The model is an agent-based microsimulation in which each individual is represented as an autonomous agent. Annual transition probabilities between health states were derived from the above sources and implemented as time-dependent functions. Monte Carlo simulation (1,000 iterations) was used to propagate parameter uncertainty, with 95% uncertainty intervals calculated from the 2.5^th^ and 97.5^th^ percentiles of the simulated outputs.

### Incidence and prevalence of ALD

Estimates of ALD incidence were derived from longitudinal studies and GBD 2021 data for China ^[1]^. To project future incidence, we fit linear regression models corresponding to 3 different age groups with age-specific incidence as a dependent variable and the year as an independent variable (Tables S2-S4). We then estimated age-specific incidence rates, using calibration with prevalence from 2000 through 2018 as targets. In the base case, we assumed ALD incidence would increase until 2030 before stabilizing and varied this assumption in sensitivity analyses.

To populate the model with the prevalence of ALD in 2020, the starting point of our simulation, we estimated the prevalence of ALD in 2018 using data from the China Kadoorie Biobank and national health surveys ^[26]^ and conducted back calculations to determine the prevalence in 2020 (Supplementary Methods, Tables S5 and S6, and Figure S1).

### Progression of ALD and proportion of ASH

To estimate transition probabilities among ALD-related health states, we extracted data from published meta-analyses of paired biopsy studies and reported the incidence of progression and regression by fibrosis stage (F0-F4) and ASH status. We estimated the rate of resolving simple steatosis to be half the incidence, based on these meta-analyses. Because published studies did not report development and resolution of ASH by fibrosis stage, we initially applied uniform rates from the meta-analyses for all fibrosis stages and then calibrated them using the proportion of ASH among ALD from 2000 through 2018 as targets. Transition from cirrhosis (F4) to DC was independent of ASH status ^[27]^.

In biopsy-based studies of patients with alcohol use disorder, the proportion of ASH ranged from 39% to 93%. Population-level data on ASH prevalence in China are limited, with no precise estimates available for Chinese adults. However, overall ALD prevalence is approximately 4.5%-4.8%, suggesting ASH affects a subset of those with ALD. There are no data on the proportion of ASH in the year 2000 or trends over time, highlighting a key gap in longitudinal epidemiological studies ^[28, 29]^. We developed an ASH prediction model using a subset of the China Kadoorie Biobank. Subsequently, we employed the prediction model and national survey data to estimate the ASH proportion among patients with ALD in 2018 and assess its temporal change. Additionally, we performed back calculations to determine the ASH proportion in year 2020 (Supplementary Methods and Tables S7-S9).

### HCC and LT

In our model, we allowed transition from F3 or F4 to HCC based on pooled longitudinal studies and meta-analyses of ALD cohorts, with an annual incidence rate of HCC among patients with advanced fibrosis or cirrhosis estimated at 1% to 4% (Table S10) ^[30–32]^. Although people with DC can develop HCC, many cases arise in compensated cirrhosis without prior decompensation, and the observed incidence in decompensated stages may appear lower due to competing risks of mortality. We assumed the transition from DC to HCC was half that from F4 to HCC in the base-case analysis and varied it in sensitivity analyses to assess robustness ^[33–35]^.

For LT, we modeled the annual probability of receiving a transplant among patients with DC or HCC based on Chinese national transplant registry data and global trends, with rates ranging from 0.1% to 0.5% annually, reflecting limited donor availability and regional disparities in access (Table S11) ^[36]^. Post-LT states included survival with potential complications, calibrated to 1-year and 5-year survival rates from cohort studies (90% and 75%, respectively) ^[37]^.

### Mortality

Mortality in the model included liver-related deaths from DC, HCC, and post-LT complications, as well as all-cause background mortality (i.e., mortality rates in the general population without ALD). Annual mortality from DC was estimated at 15% to 20% in the first year of decompensation, decreasing to 10% to 15% thereafter, based on prospective cohort studies and meta-analyses of alcohol-associated liver disease patients (Table S12) ^[38, 39]^. For HCC, mortality rates varied by time since diagnosis: 40% to 60% in the first year, 20% to 40% in subsequent years, derived from Surveillance, Epidemiology, and End Results (SEER) data adapted to Chinese registries and adjusted for alcohol-associated liver disease-specific risks ^[6, 40]^. Post-LT mortality was age-dependent, with rates of 5% to 10% in the first year and 2% to 5% annually thereafter, sourced from international transplant databases and Chinese studies ^[41]^.

Baseline all-cause mortality was incorporated using age- and sex-specific rates from Chinese national life tables (2020-2050 projections), with a hazard ratio (HR) of 1.2 to 1.5 for individuals with ALD compared to those without, to account for excess non-liver mortality (e.g., cardiovascular risks) associated with chronic alcohol use (Table S13) ^[42, 43]^. Liver-related mortality was calculated as the excess over background rates.

### Disability-adjusted life years (DALYs)

DALYs were calculated as the sum of years of life lost (YLL) due to premature mortality and years lived with disability (YLD) ^[44]^. YLL were computed for each simulated death by multiplying the number of deaths in each age-sex group by the remaining life expectancy at the age of death, using age- and sex-specific life tables from the UNWPP 2024 medium variant. YLD were calculated by multiplying the person-years lived in each health state (ASL, ASH, fibrosis stages F0-F4, DC, HCC, and post-LT) by the corresponding disability weight for that state. Disability weights were derived from the GBD 2021 for ALD states: compensated cirrhosis 0.178, decompensated cirrhosis 0.432, HCC 0.496, and post-LT 0.10-0.25, depending on complications ^[45]^. Only liver-related DALYs were included in the primary analysis. Non-liver excess mortality was accounted for in the baseline mortality HR but not assigned additional disability weights. All DALYs were aggregated across the simulated cohort and reported annually from 2020 to 2050 with 95% uncertainty intervals derived from 1,000 Monte Carlo simulations.

### Statistical analysis

All analyses were performed using R version 4.5.3 (R Project for Statistical Computing). The model was run with 1,000 Monte Carlo simulations to propagate uncertainty in input parameters, generating 95% UI as the 2.5^th^ and 97.5^th^ percentiles of the simulation outputs. Parameter uncertainty was sampled from beta distributions for probabilities, gamma distributions for rates, and log-normal distributions for HRs, based on reported variances or assumed 20% coefficients of variation when not available. Sensitivity analyses included one-way variations (±20% for key parameters like progression rates and incidence trends) and scenario analyses for alcohol policy interventions.

## Results

### Model calibration and validation

The model simulated 5,678,912 individuals (mean age, 36.2 years; 51.2% male). The state-transition model was calibrated using baseline prevalence estimates from meta-analyses, which reported a pooled prevalence of ALD in China of 23.64% (95% UI 18.41%-29.20%) among heavy drinkers, based on histological data from multiple studies ^[24]^. Progression rates were informed by systematic reviews, with annual transitions as follows: from steatosis to cirrhosis at 3% (95% UI 2%-4%), from steatohepatitis to cirrhosis at 10% (95% UI 6%-17%), and from fibrosis to cirrhosis at 8% (95% UI 3%-19%) ^[46]^. HCC incidence rates were calibrated to 1% to 4% annually in advanced fibrosis or cirrhosis, drawing from pooled longitudinal data ^[30]^. Mortality rates were calibrated against GBD 2021 data, which estimated approximately 50,000 alcohol attributable cirrhosis deaths in Chinese adults under the age of 70 ^[23]^. The model validated well, reproducing 48,500 deaths in the base year (95% UI 40,000-57,000) and 42,000 HCC cases (95% UI 35,000-49,000), with sensitivity analyses confirming robustness to variations in drinking patterns from China Health and Retirement Longitudinal Study (CHARLS) waves (Figure S1) ^[26]^. Transition probabilities used in the model are detailed in Table S14.

### Projected prevalence of ALD

Under the business-as-usual scenario, assuming continued increases in alcohol consumption (from 7.1 L to 11.2 L per capita annually, per WHO trends), the prevalence of ALD among Chinese adults is projected to rise from 4.8% in 2020 to 8.5% in 2050 (95% UI 6.2%-10.8%) (Table 1 and Figure 2). This corresponds to an increase in affected individuals from approximately 55 million in 2020 to 94 million in 2050, driven by population aging (UNWPP) and rising hazardous drinking rates (e.g. 33% of men drink regularly at 286 g/week in China) ^[47]^. Prevalence is highest in advanced stages by 2050: cirrhosis at 2.6% (95% UI 1.9%-3.3%), steatohepatitis at 2.0% (95% UI 1.4%-2.6%). Subgroup analysis shows stark sex differences: male prevalence reaches 12.3% (95% UI 9.0%-15.6%) by 2050, vs. 4.7% (95% UI 3.4%-6.0%) in females, reflecting CHARLS data on low female drinking (2%) (Table S1) ^[26, 29]^.

**Figure 2.**
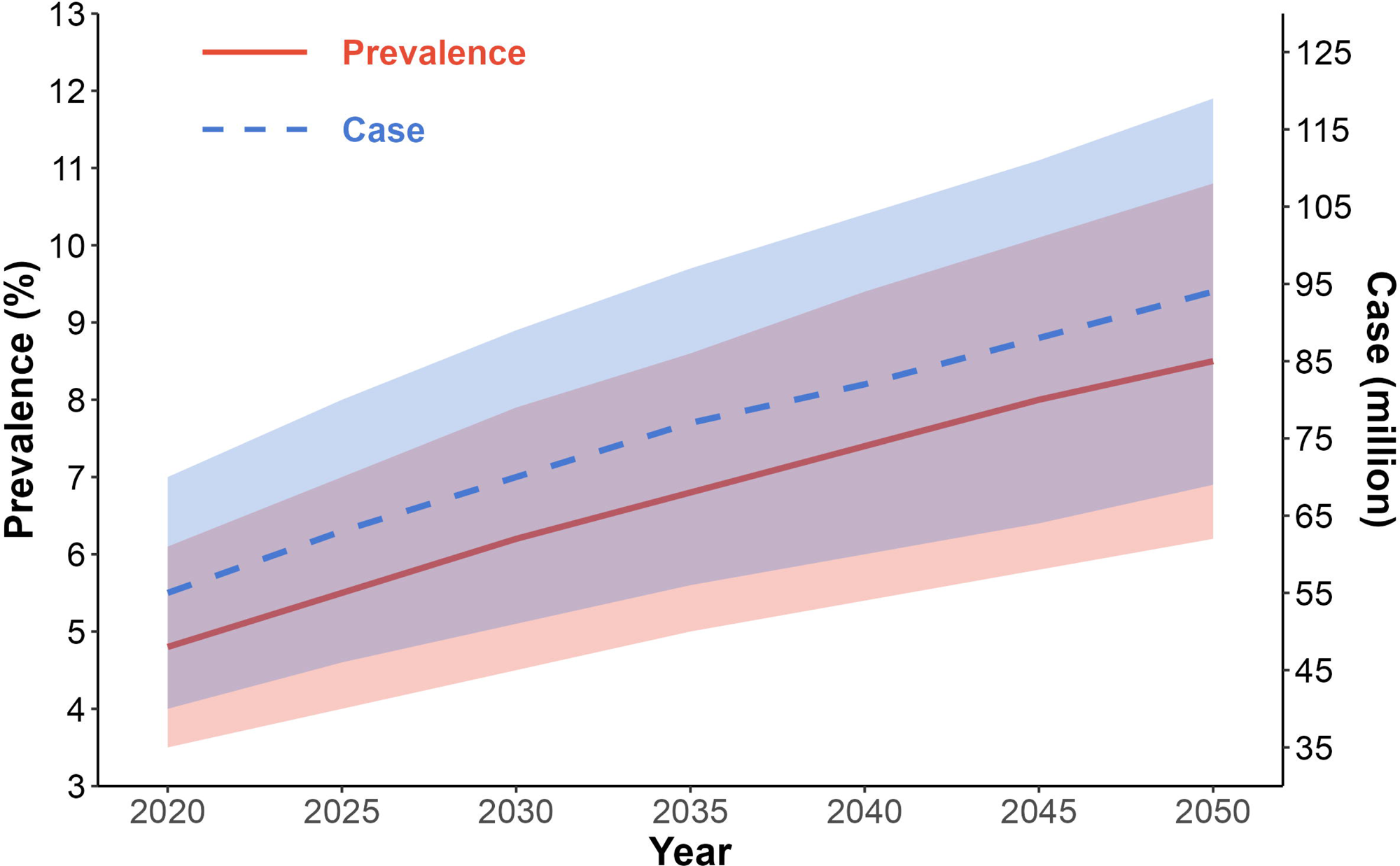
Projected prevalence of alcohol-associated liver disease (ALD) among Chinese adults, 2020-2050, under the business-as-usual scenario. Solid blue line indicates point estimates; shaded blue area represents 95% uncertainty intervals.

**Table 1.** Projected prevalence of ALD in Chinese adults, 2020-2050.

| Year | Prevalence (%) (95% UI) | Number of cases (millions) (95% UI) |
| --- | --- | --- |
| 2020 | 4.8 (3.5-6.1) | 55 (40-70) |
| 2030 | 6.2 (4.5-7.9) | 70 (51-89) |
| 2040 | 7.4 (5.4-9.4) | 82 (60-104) |
| 2050 | 8.5 (6.2-10.8) | 94 (69-119) |

### Projected incidence of HCC and LT

Annual incident HCC cases are projected to rise from 20,500 (95% UI 15,000-26,000) in 2020-2025 to 45,200 (95% UI 33,000-57,400) by 2046-2050, reflecting higher progression in heavy drinkers (Figure 2). LT demand is forecasted to increase from 2,300 (95% UI 1,700-2,900) annually in 2020-2025 to 9,800 (95% UI 7,200-12,400) by 2046-2050, based on calibrated rates from limited donor availability and rising end-stage disease (Figure 3 and Table S11). These increases are most pronounced in males (70% of excess HCC cases) and urban populations (60% of excess LT demand), consistent with epidemiological patterns ^[47]^.

**Figure 3.**
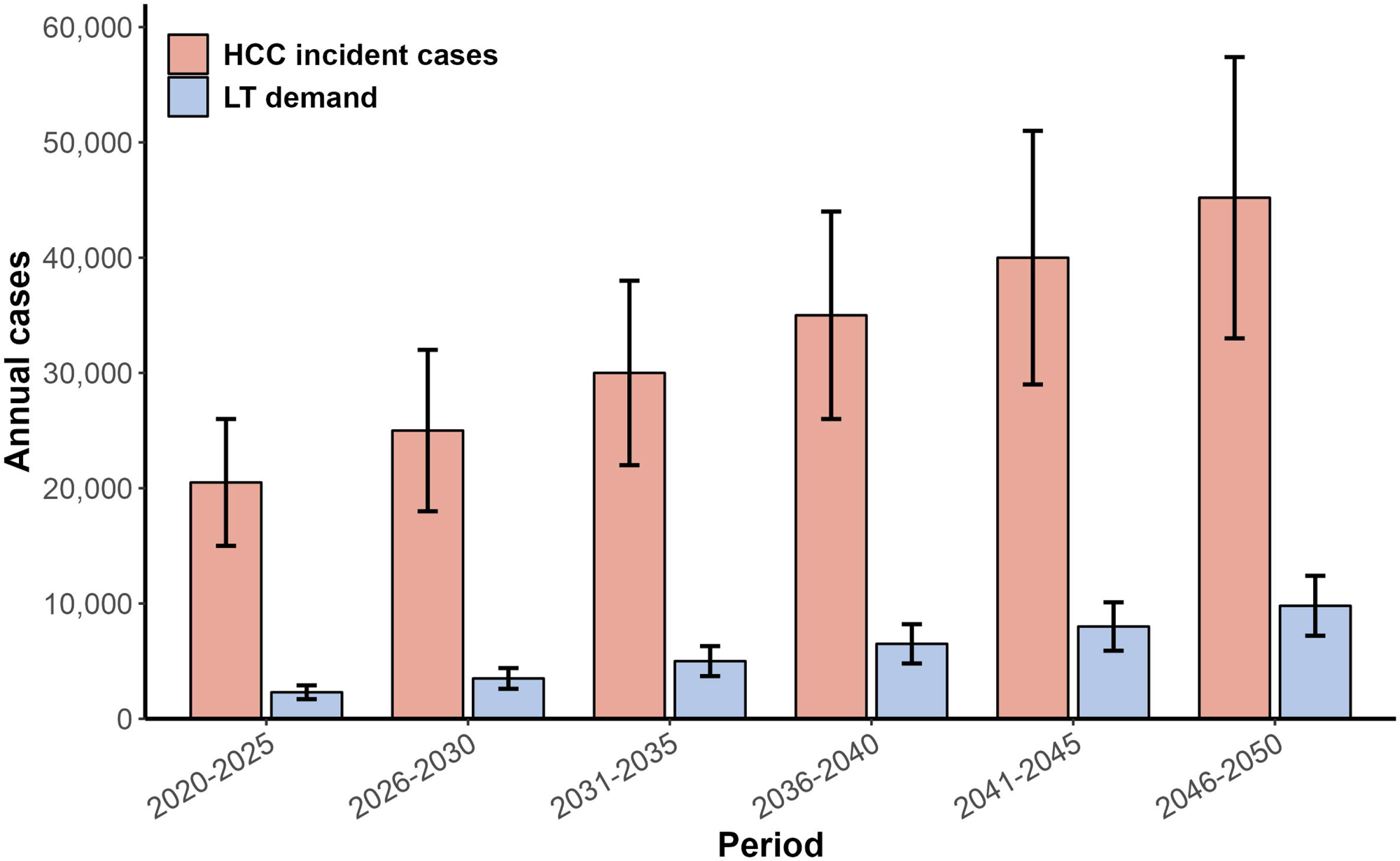
Projected annual incident cases of hepatocellular carcinoma (HCC) and liver transplants (LT), 2020-2050. Error bars show 95% uncertainty intervals.

### Projected mortality and DALYs

ALD-attributable deaths are projected to increase from 50,000 in 2020 to 85,000 in 2050 (95% UI 70,000-100,000), with liver-specific causes (e.g., cirrhosis complications) accounting for approximately 75% of excess mortality (Figure 3). DALYs lost due to ALD are expected to rise from 1.5 million in 2020 to 2.6 million in 2050 (95% UI 2.1-3.1 million) (Figure 4), incorporating both premature deaths and years lived with disability from progressive stages ^[23]^. These projections align with observed trends where alcohol contributes to multiple disease risks, including cirrhosis and gout ^[47]^.

**Figure 4.**
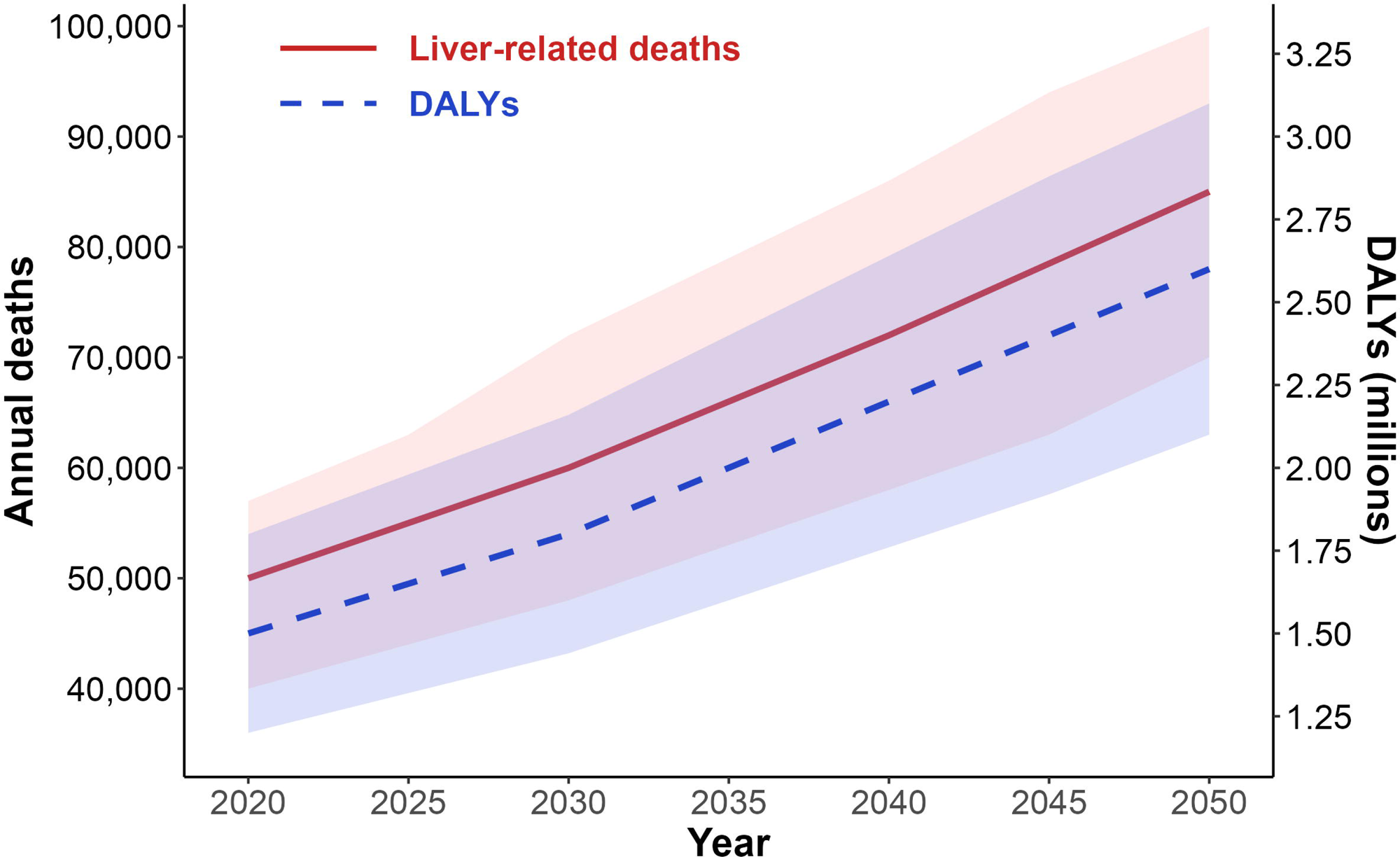
Projected annual liver-related deaths (left Y-axis) and disability-adjusted life years (DALYs) lost (right Y-axis) due to alcohol-associated liver disease (ALD), 2020-2050. Shaded areas indicate 95% uncertainty intervals.

### Sensitivity and subgroup analyses

Sensitivity analyses varying progression rates (±20%) yielded prevalence estimates of 7.2%-9.8% by 2050, confirming model stability (Figure S2). Subgroup projections highlight urban-rural disparities (urban prevalence 9.2% vs. rural 7.8% by 2050) and age effects: prevalence peaks at 10.1% in 50-69-year-olds (Table S15). Genetic evidence supports causality, with alcohol metabolism variants linked to higher risks of cirrhosis (HR 2.30, 95% CI 1.58-3.35) and gout (HR 2.33, 95% CI 1.49-3.62), reinforcing projections (Table S15) ^[47]^. Scenario analyses evaluating alcohol policy interventions (e.g., 10%–30% reduction in heavy drinking) projected that such measures could avert 25-40% of the forecasted ALD cases, HCC incidence, and liver-related deaths by 2050 (Table 2).

**Table 2.** Projected impact of alcohol policy interventions on ALD burden in Chinese adults in 2050.

Chinese adults in 2050
| Scenario | ALD cases<br>(millions) | Annual incident<br>HCC cases | Annual<br>liver-related<br>deaths | DALYs<br>(millions) |
| --- | --- | --- | --- | --- |
| Base-case<br>(business-as-usual) | 94 | 45,200 | 85,000 | 2.6 |
| 10% reduction in heavy<br>drinking | 82 | 39,500 | 74,000 | 2.3 |
| 20% reduction in heavy<br>drinking | 71 | 34,200 | 64,000 | 2.0 |
| 30% reduction in heavy<br>drinking | 59 | 28,400 | 53,000 | 1.7 |

## Discussion

This decision analytical modeling study projects a substantial and sustained increase clinical burden of ALD among Chinese adults between 2020 to 2050 under a business-as-usual scenario. In the absence of strengthened alcohol control policies, the prevalence of ALD is forecasted to increase from 4.8% (approximately 55 million individuals) in 2020 to 8.5% (approximately 94 million) by 2050, with corresponding rises in cirrhosis (to 2.6%) and steatohepatitis (to 2.0%). Annual incident HCC cases are projected to more than double (from 20,500 to 45,200), liver transplant demand to quadruple (from 2,300 to 9,800), and liver-related deaths to rise from 50,000 to 85,000, accounting for an increasing share of adult all-cause mortality (from 1.2% to 3.1%). DALYs lost due to ALD are expected to increase from 1.5 million to 2.6 million over the same period ^[23, 24, 48]^. These projections confirm that ALD is poised to emerge as a dominant cause of liver-related morbidity, mortality, and health system demand in China, particularly as control of viral hepatitis continues to improve.

Three interconnected and mutually reinforcing drivers underpin these projected trends. First, per capita alcohol consumption in China has risen markedly over recent decades and is expected to keep increasing without strengthened policy intervention, raising population-wide exposure to hazardous drinking. Second, population aging will shift the demographic structure toward older age groups, who carry higher risks of ALD progression, advanced fibrosis, and HCC. Third, approximately 33% of men engage in regular heavy drinking at a mean intake of 286 g per week, and such socially normalized heavy episodic drinking creates a large high-risk pool for progressive liver disease. Male ALD prevalence is projected to reach 12.3% by 2050, compared with only 4.7% in women, a disparity consistent with national surveys showing low female drinking prevalence (around 2%) ^[26, 47]^. The burden will concentrate disproportionately among urban men aged 50-69 years, representing a priority group for targeted intervention ^[29]^.

Our results align closely with national and global estimates showing that alcohol-attributable liver mortality already exceeds 50,000 deaths annually among adults younger than 70 years in China ^[23, 49]^. The projected growth in disease burden is also consistent with global burden of disease reports highlighting the rising contribution of alcohol to chronic liver disease. Notably, our findings contrast sharply with trends in many Western countries, where comprehensive alcohol policy packages have stabilized or reduced alcohol-related liver disease burdens. China’s trajectory reflects unique sociocultural factors, including the normalization of heavy drinking in business, social, and ceremonial settings, which underscores the need for culturally tailored alcohol control strategies rather than one-size-fits-all approaches ^[46, 50]^.

This study has notable strengths that enhance the robustness and policy relevance of its findings. First, we employed an extra-large simulated cohort of 5,678,912 individuals, enabling stable estimation of rare events such as HCC and LT. Second, the model fully incorporated China-specific demographic trends and real-world drinking patterns, which greatly improves national representativeness. Third, we explicitly projected HCC incidence and LT clinical need, which are critical outcomes frequently overlooked in previous burden assessments. Finally, our agent-based microsimulation framework captures individual-level heterogeneity, yielding far more granular and reliable projections than traditional aggregate cohort models ^[51]^.

Several limitations should be acknowledged. First, disease progression rates were derived mainly from hospital-based or biopsy-confirmed cohorts, which may overestimate risks in the general population ^[24]^. Second, although alcohol consumption trends were informed by WHO reports and longitudinal surveys, granular grams-per-day exposure data at the individual level were limited, introducing uncertainty in incidence projections ^[29]^. Third, synergistic interactions between ALD and metabolic comorbidities were not fully incorporated despite their potential to accelerate liver damage. Fourth, data on alcoholic steatohepatitis proportions and long-term trends before 2000 remain sparse, necessitating calibration that adds to uncertainty intervals. Finally, although the model was validated against GBD mortality and HCC estimates, future refinements could integrate regional variations from ongoing national surveillance.

Without urgent and strengthened alcohol control policies, the projected surge in HCC, DC, and end stage liver disease will place unprecedented strain on China’s health systems, particularly tertiary medical center that provide liver transplantation and cancer treatment. Evidence-based interventions could avert an estimated 30-40% of the predicted disease burden by 2050. These interventions include increased alcohol excise taxation, restrictions on advertising and marketing, reduced physical availability, and integration of ALD screening into routine primary care. Multisectoral collaboration across hepatology, endocrinology, primary care, public health, and health policy will be essential to translate these findings into actionable, sustainable, and equitable prevention strategies ^[52]^.

## Conclusions

ALD will become a defining public health and clinical challenge in China over the next 30 years unless effective, comprehensive, and culturally appropriate alcohol control measures are implemented urgently. Sustained policy action is critical to mitigate preventable liver-related deaths, curb rising HCC incidence, reduce the growing demand for liver transplantation, and alleviate the population-level burden of life-years lost to disability and premature mortality. These findings offer timely evidence to support policymakers in prioritizing alcohol control and liver disease prevention within national health strategies, while helping China progress toward WHO targets for reducing harmful alcohol use.

## Data Availability

All data produced in the present study are available upon reasonable request to the authors

## List of Abbreviations

ALD: Alcohol-associated liver disease
ASH: alcoholic steatohepatitis
ASL: alcohol-associated steatotic liver
DALYs: disability-adjusted life years
DC: decompensated cirrhosis
GBD: Global Burden of Disease
HCC: hepatocellular carcinoma
HR: hazard ratio
LT: liver transplantation
MASLD: metabolic dysfunction-associated steatotic liver disease
SEER: Surveillance, Epidemiology, and End Results
STRESS: Strengthening the Reporting of Empirical Simulation Studies
YLD: years lived with disability
YLL: years of life lost
UI: uncertainty intervals
UNWPP: UN World Population Prospects

## Conflict of interest

Nothing to declare.

## Author contributions

All authors approve the final version of the manuscript, including the authorship list, and agree to be accountable for all aspects of the work in ensuring that questions related to the accuracy or integrity of any part of the work are appro priately investigated and resolved. All authors have read and approved the final version of the manuscript for submission. Qingfeng Niu: Study concept and design; manuscript writing, editing, and critical review. Mingzhu Su:Manuscript editing. Leshi Liang: Data acquisition and analyses. Zhaoyi Che: Data acquisition and analyses. Qiang Zhu: Manuscript critical review. Fei Wang: Study concept and design; Data acquisition and analyses; manuscript writing, editing, and critical review. Jia Xiao: Data acquisition and analyses; manuscript writing, editing, and critical review.

## Funding

This study was supported by Ministry of Science and Technology of China (MOST) National Key R&D Program (Grants 2023YFA1800800), Shenzhen Medical Research Funds (B2302007), and National Natural Science Foundation of China (U23A20401).

## Data Availability

The statistical and analytic code will be provided upon reasonable request. Interested researchers may send requests to the corresponding author.

---

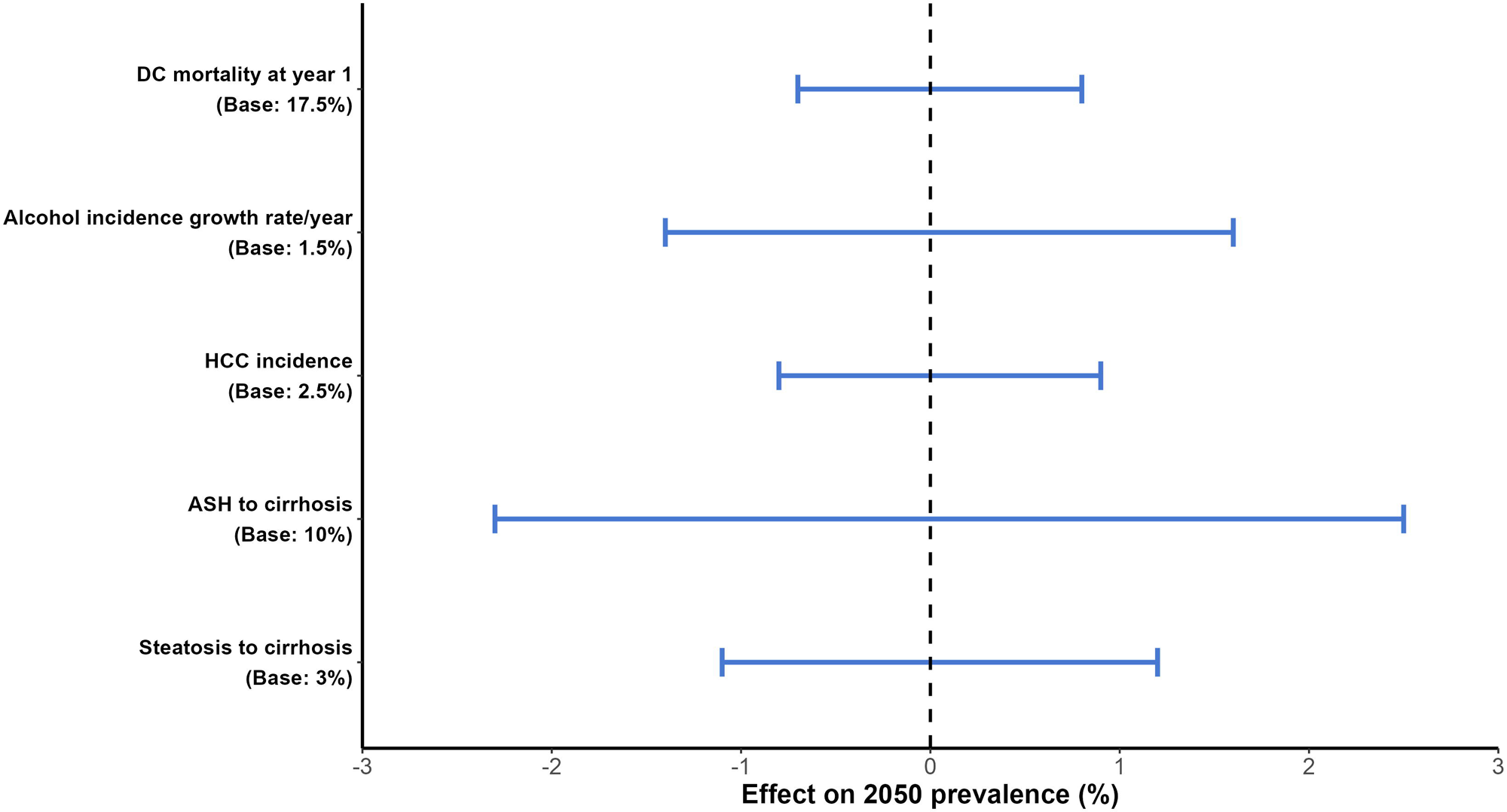

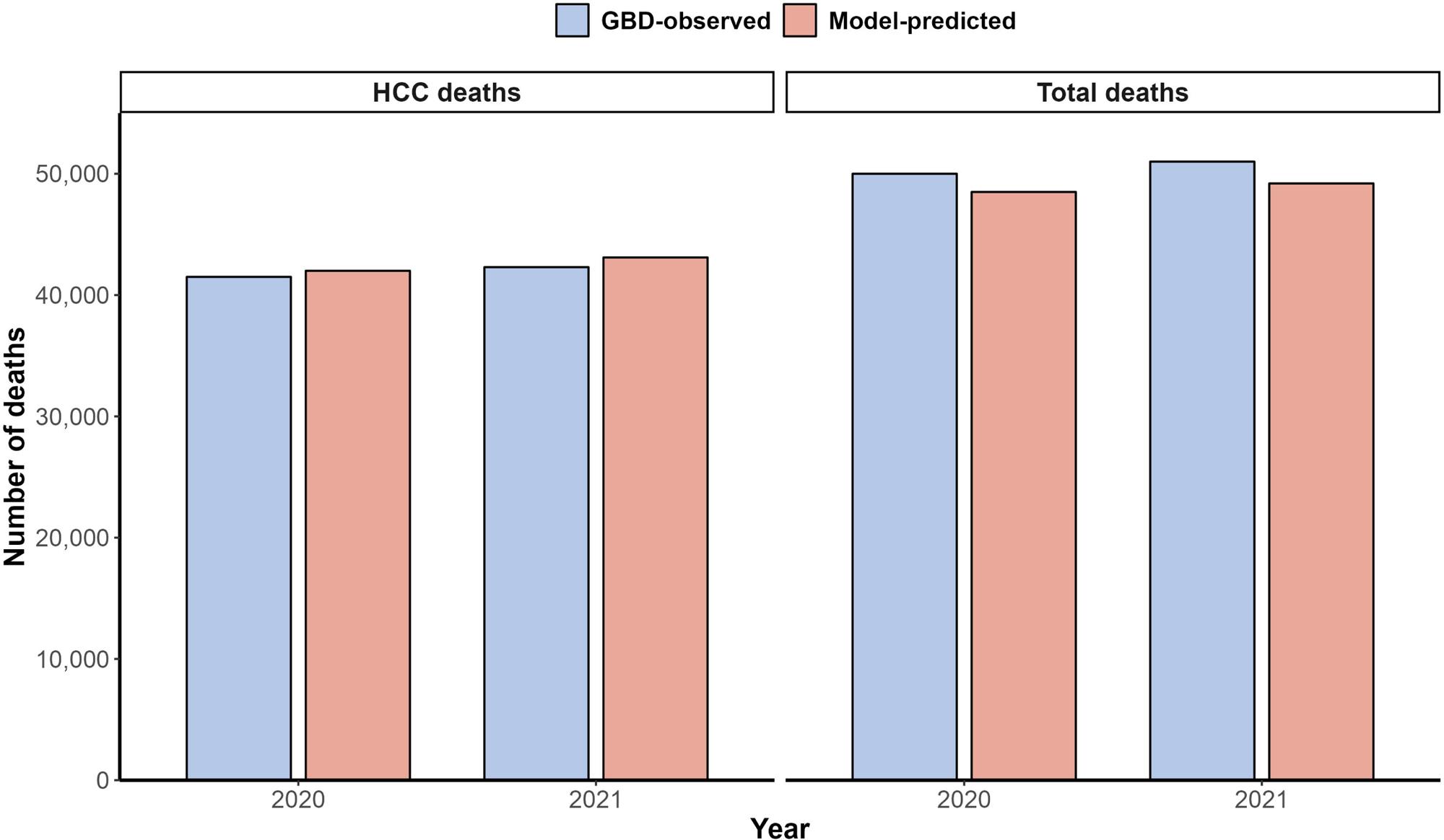

## Notes

### Competing Interest Statement

The authors have declared no competing interest.

### Author Declarations

This study will only use openly available human data that were originally located at published papers

