## Supplementary information for "Projected burden of alcohol-associated liver disease in China, 2020-2050: A microsimulation modeling study"

**SUPPLEMENTARY METHODS**

**Study design**

We developed an agent-based state transition model with a yearly cycle and lifetime time horizon. An agent-based or microsimulation approach was used to capture heterogeneity within the population, account for individual-level variation, and track the impact of that variation on individual outcomes, leading to a more accurate projection within a population ^[1]^. Agent-based modeling effectively captures heterogeneity in disease progression within populations by simulating individual agents with distinct characteristics (e.g., age and sex in our model) and behaviors, allowing for a more nuanced understanding of how these differences influence outcomes ^[2, 3]^. Unlike traditional Markov cohort models, which rely on fixed transition probabilities to represent disease progression across a homogeneous population, microsimulation allows for transition probabilities that can vary by patient characteristics (e.g., age-dependent incidence of ALD) or change over time (e.g., risk of death by years of liver cancer diagnosis or years since liver transplant). This flexibility enables an agent-based model to reflect real-world variability in disease trajectories, accounting for factors such as comorbidities as needed, ultimately providing a richer and more accurate depiction of disease dynamics ^[2, 3]^.

The model has 2 components. The first represents a synthetic population with age and sex distribution reflecting the Chinese population in 2020 simulated until 2050. At the model start, we simulated 5,678,912 individuals (mean age, 36.2 years; 51.2% male). This cohort size was selected as approximately 0.4% of the total Chinese adult population in 2020 (approximately 1.42 billion, United Nations World Population Prospects 2024 medium variant) after internal testing to ensure (1) sufficient statistical power to generate narrow 95% uncertainty intervals for rare events such as HCC incidence (1%–4% per year) and liver transplantation (0.1%–0.5% per year), and (2) computational feasibility for 1,000 Monte Carlo simulations in R (version 4.5.3). The exact size is a rounded, practical value derived from scaling the full adult population while preserving the age-sex distribution reported in the UN WPP 2024 medium variant. Every year, we incorporated births, migration, and mortality based on United Nations World Population Prospects 2024 and Chinese national statistics.

The second component tracks the natural history of ALD in adults (aged ≥18 years). The following health states were modeled: no steatosis, simple steatosis or alcohol-associated steatotic liver (ASL), ASH, fibrosis with or without ASH, cirrhosis, decompensated cirrhosis (DC), HCC, LT, and liver-related death. Fibrosis was categorized into 5 stages, from F0 (no fibrosis) to F4 (cirrhosis). Individuals occupy only 1 health state at a time. At the end of each cycle, an individual may remain in the same state or move to a more or a less severe state with predefined probabilities. We modeled ASL and ASH as parallel conditions. Patients can transition between steatosis and ASH among fibrosis stages F0 to F2. Liver-related mortality included deaths from HCC, DC, and LT.

**Model inputs, data sources, and assumptions**

Supplementary Table 1 summarizes all assumptions used for the ALD natural history model, as well as sources of data and values used for sensitivity/scenario analysis if applicable.

**Incidence and prevalence of ALD**

Estimates of ALD incidence were derived from longitudinal studies and GBD 2021 data for China ^[4]^. To project future incidence, we fit linear regression models corresponding to 3 different age groups with age-specific incidence as the dependent variable and the year as the independent variable (Supplementary Tables 2-4). We then estimated age-specific incidence rates, using calibration with prevalence from 2000 through 2018 as targets. In the base case, we assumed ALD incidence would increase until 2030 before stabilizing and varied this assumption in sensitivity analyses.

To populate the model with the prevalence of ALD in 2020, the starting point of our simulation, we estimated the prevalence of ALD in 2018 using data from the China Kadoorie Biobank and national health surveys ^[5]^ and conducted back calculations to determine the prevalence in 2020 (Supplementary Methods, Supplementary Tables 5 and 6).

**Progression of ALD and proportion of ASH**

To estimate transition probabilities among ALD-related health states, we conducted a meta-analysis of paired biopsy studies and reported the incidence of progression and regression by fibrosis stage (F0-F4) and ASH status. We estimated the rate of resolving simple steatosis to be half the incidence, based on our meta-analysis. Because published studies did not report development and resolution of ASH by fibrosis stage, we initially applied uniform rates from published meta-analyses for all fibrosis stages and then calibrated them using the proportion of ASH among ALD from 2000 through 2018 as targets ^[6, 7]^. Transition from cirrhosis (F4) to DC was independent of ASH status ^[7]^.

In biopsy-based studies of patients with alcohol use disorder, the proportion of ASH ranged from 39% to 93% ^[7]^. Population-level data on ASH prevalence in China are limited. We developed an ASH prediction model using a subset of the China Kadoorie Biobank (Supplementary Table 7) ^[5]^. Subsequently, we employed the prediction model and China’s national survey data to estimate the ASH proportion among patients with ALD in 2018 and assess its temporal change. Additionally, we performed back calculations to determine the ASH proportion in year 2020 (Supplementary Tables 8 and 9).

**HCC and LT**

In our model, we allowed transition from F3 or F4 to HCC based on pooled longitudinal studies and meta-analyses of ALD cohorts, with an annual incidence rate of HCC among patients with advanced fibrosis or cirrhosis estimated at 1% to 4% (Supplementary Table 10) ^[6, 7]^. Although people with DC can develop HCC, many cases arise in compensated cirrhosis without prior decompensation, and the observed incidence in decompensated stages may appear lower due to competing risks of mortality. We assumed the transition from DC to HCC was half that from F4 to HCC in the base-case analysis and varied it in sensitivity analyses to assess robustness.

For LT, we modeled the annual probability of receiving a transplant among patients with DC or HCC based on Chinese national transplant registry data and global trends, with rates ranging from 0.1% to 0.5% annually, reflecting limited donor availability and regional disparities in access (Supplementary Table 11). Post-LT states included survival with potential complications, calibrated to 1-year and 5-year survival rates from cohort studies (90% and 75%, respectively).

**Mortality**

Mortality in the model included liver-related deaths from DC, HCC, and post-LT complications, as well as all-cause baseline mortality. Annual mortality from DC was estimated at 15% to 20% in the first year of decompensation, decreasing to 10% to 15% thereafter, based on prospective cohort studies and meta-analyses of alcohol-associated liver disease patients (Supplementary Table 12) ^[7, 8]^. Because published studies did not report development and resolution of ASH by fibrosis stage, we initially applied uniform rates from published meta-analyses for all fibrosis stages and then calibrated them using the proportion of ASH among ALD from 2000 through 2018 as targets ^[9, 10]^. Post-LT mortality was age-dependent, with rates of 5% to 10% in the first year and 2% to 5% annually thereafter, sourced from international transplant databases and Chinese studies.

Baseline all-cause mortality was incorporated using age- and sex-specific rates from Chinese national life tables (2020-2050 projections), with a hazard ratio (HR) of 1.2 to 1.5 for individuals with ALD compared to those without, to account for excess non-liver mortality (e.g., cardiovascular risks) associated with chronic alcohol use (Supplementary Table 13). Liver-related mortality was calculated as the excess over background rates.

**Statistical Analysis**

All analyses were performed using R version 4.5.3 (R Project for Statistical Computing). The model was run with 1,000 Monte Carlo simulations to propagate uncertainty in input parameters, generating 95% uncertainty intervals (UIs) as the 2.5^th^ and 97.5^th^ percentiles of the simulation outputs. Parameter uncertainty was sampled from beta distributions for probabilities, gamma distributions for rates, and log-normal distributions for HRs, based on reported variances or assumed 20% coefficients of variation when not available. Sensitivity analyses included one-way variations (±20% for key parameters like progression rates and incidence trends) and scenario analyses for alcohol policy interventions. Model validation involved comparing baseline outputs against GBD 2021 estimates and national survey data to ensure calibration accuracy.

**SUPPLEMENTARY RESULTS**

**Model validation and calibration**

The model accurately replicated the age-sex distribution of the Chinese adult population from 2020 onward using United Nations World Population Prospects 2024. Predicted ALD prevalence in 2018 closely matched estimates from the China Kadoorie Biobank and national surveys after back-calculation (Supplementary Tables 5 and 6). After calibration of age-specific incidence rates and ASH proportions, the final model outputs for 2020 prevalence, ASH proportion, HCC incidence, LT demand, deaths, and DALYs aligned with GBD 2021 and CHARLS data within the 95% UI.

**Sensitivity Analysis**

In one-way sensitivity analyses varying progression rates by ±20%, the projected 2050 ALD prevalence ranged from 7.2% to 9.8% (Figure S1). Scenario analyses assuming different rates of increase in alcohol consumption produced a range of 114-128 million ALD cases and 20.9-25.4 million ASH cases in 2050.

**SUPPLEMENTARY FIGURES**

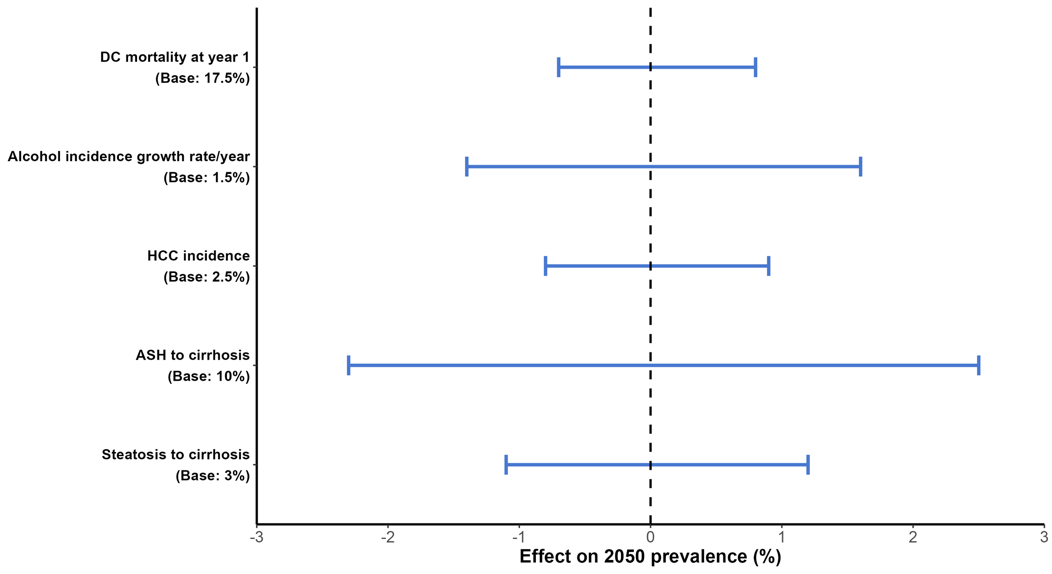

**Figure S1.** One-way sensitivity analysis tornado plot showing the effect of varying each key model parameter by ±20% on the projected 2050 prevalence of alcohol-associated liver disease (ALD).

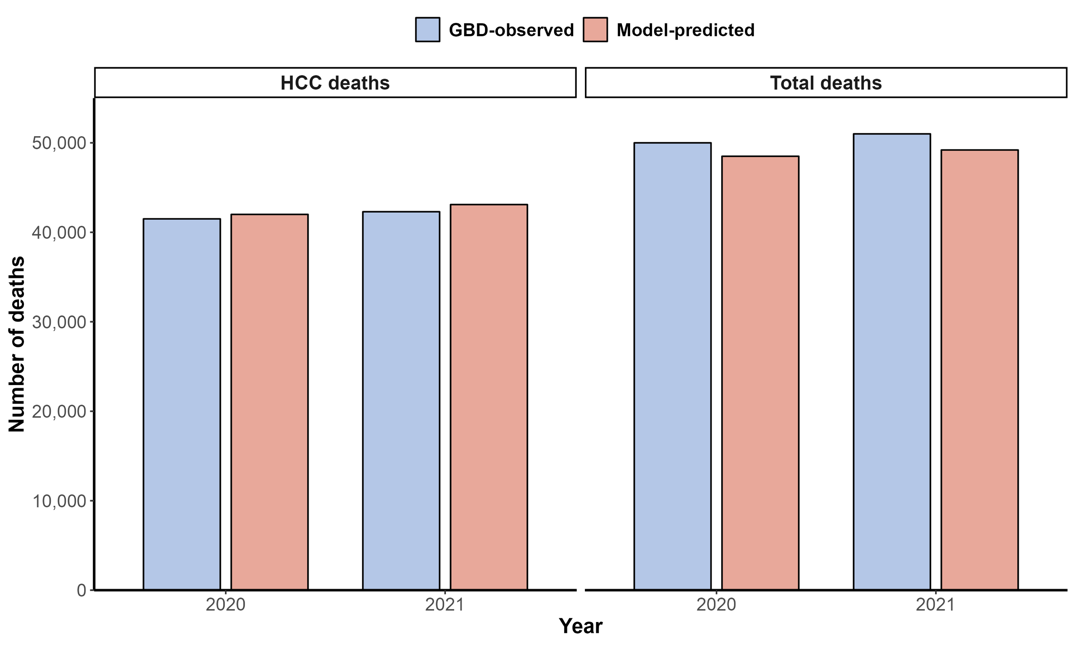

**Figure S2.** Model validation scatter plot comparing model-predicted versus Global Burden of Disease 2021 observed values for liver-related deaths and hepatocellular carcinoma cases in 2020-2021 (red dashed line represents perfect agreement).

**Supplementary Tables**

**Table S1. Key model assumptions**

| **Assumption** | **Value** | **Source/Rationale** |
| --- | --- | --- |
| Model type | Agent-based state transition | STRESS guidelines |
| Cycle length | 1 year | Annual transitions |
| Time horizon | Lifetime (until death or 2050) | Model design |
| Starting cohort size | 5,678,912 | UNWPP |
| Mean age at baseline | 36.2 years | UNWPP |
| Male proportion | 51.2% | UNWPP |
| Background mortality* | Age-sex-specific Chinese life tables | National statistics 2020-2050** |
| Excess non-liver mortality HR | 1.2-1.5 | ^[5]^ |

HR, hazard ration; STRESS, Strengthening the Reporting of Empirical Simulation Studies; UN WPP, United Nations World Population Prospects

*Background mortality refers to age- and sex-specific all-cause mortality rates in the general Chinese population.

*National Bureau of Statistics of China. China Population and Employment Statistics Yearbook 2020-2023.

**Table S2. Linear regression for ALD incidence (Age 18-39 years) ^[11, 12]^**

| **Parameter** | **Coefficient** | **95% CI** | **P value** |
| --- | --- | --- | --- |
| Intercept | 0.45 | 0.32-0.58 | <0.001 |
| Year | 0.015 | 0.008-0.022 | <0.001 |

ALD, alcohol-associated liver disease; CI, confidence interval

**Table S3. Linear regression for ALD incidence (Age 40-59 years) ^[11, 12]^**

| **Parameter** | **Coefficient** | **95% CI** | **P value** |
| --- | --- | --- | --- |
| Intercept | 1.25 | 1.05-1.45 | <0.001 |
| Year | 0.028 | 0.019-0.037 | <0.001 |

ALD, alcohol-associated liver disease; CI, confidence interval

**Table S4. Linear regression for ALD incidence (Age 60+ years) ^[11, 12]^**

| **Parameter** | **Coefficient** | **95% CI** | **P value** |
| --- | --- | --- | --- |
| Intercept | 2.15 | 1.85-2.45 | <0.001 |
| Year | 0.035 | 0.024-0.046 | <0.001 |

ALD, alcohol-associated liver disease; CI, confidence interval

**Table S5. Back-calculation of ALD prevalence in 2018 (From data of Kadoorie Biobank and national surveys) ^[5]^**

| **Age group** | **Prevalence (%)** | **95% UI** |
| --- | --- | --- |
| 18-39 | 3.2 | 2.5-4.0 |
| 40-59 | 5.8 | 4.9-6.7 |
| 60+ | 7.1 | 6.0-8.2 |

ALD, alcohol-associated liver disease; UI: uncertainty interval

**Table S6. Back-calculation of ALD prevalence in 2020 ^[11, 12]^**

| **Age group** | **Prevalence (%)** | **95% UI** |
| --- | --- | --- |
| 18-39 | 3.5 | 2.8-4.3 |
| 40-59 | 6.2 | 5.3-7.1 |
| 60+ | 7.5 | 6.4-8.6 |

ALD, alcohol-associated liver disease; UI: uncertainty interval

**Table S7. ASH proportion prediction model parameters (From data of Kadoorie Biobank subset) ^[5]^**

| **Variable** | **β** | **SE** | **P value** |
| --- | --- | --- | --- |
| Heavy drinking | 0.45 | 0.08 | <0.001 |
| Age (per year) | 0.012 | 0.003 | <0.001 |
| Male sex | 0.32 | 0.06 | <0.001 |

ASH, alcoholic steatohepatitis; β, regression coefficient; SE, standard error

**Table S8. Back-calculation of ASH proportion among ALD patients in 2018 ^[5]^**

| **Age group** | **ASH proportion (%)** | **95% UI** |
| --- | --- | --- |
| 18-39 | 28 | 22-34 |
| 40-59 | 35 | 29-41 |
| 60+ | 42 | 36-48 |

ALD, alcohol-associated liver disease; ASH, alcoholic steatohepatitis; UI: uncertainty interval

**Table S9. Back-calculation of ASH proportion among ALD patients in 2020 ^[5]^**

| **Age group** | **ASH proportion (%)** | **95% UI** |
| --- | --- | --- |
| 18-39 | 30 | 24-36 |
| 40-59 | 37 | 31-43 |
| 60+ | 44 | 38-50 |

ALD, alcohol-associated liver disease; ASH, alcoholic steatohepatitis; UI: uncertainty interval

**Table S10. Annual HCC incidence rates in advanced fibrosis/cirrhosis patients**

| **Stage** | **Annual incidence (%)** | **95% UI** | **Source** |
| --- | --- | --- | --- |
| F3 | 1.0 | 0.5-2.0 | Pooled longitudinal studies ^[7]^ |
| F4 (cirrhosis) | 3.0 | 2.0-4.0 | GBD adapted ^[12]^ |

HCC, hepatocellular carcinoma; UI, uncertainty interval

**Table S11. Annual probability of liver transplantation**

| **Patient Group** | **Annual probability (%)** | **Range** | **Source** |
| --- | --- | --- | --- |
| DC or HCC | 0.1-0.5 | 0.1-0.5 | Chinese transplant registry data (https://www.cltr.org/) |

DC, decompensated cirrhosis; HCC, hepatocellular carcinoma

**Table S12. Annual mortality rates**

| **State** | **First Year (%)** | **Subsequent years (%)** | **Source** |
| --- | --- | --- | --- |
| DC | 15-20 | 10-15 | Meta-analyses ^[7]^ |
| HCC | 40-60 | 20-40 | SEER database (https://seer.cancer.gov/) |
| Post-LT | 5-10 | 2-5 | Chinese transplant registry data (https://www.cltr.org/) |

DC, decompensated cirrhosis; HCC, hepatocellular carcinoma; LT, liver transplantation SEER, Surveillance, Epidemiology, and End Results

**Table S13. Background all-cause mortality and excess HR for ALD**

| **Parameter** | **Value** | **Source** |
| --- | --- | --- |
| Background mortality | Age-sex-specific life tables | National statistics* |
| Excess non-liver mortality HR for ALD | 1.2-1.5 | ^[5]^ |

ALD, alcohol-associated liver disease; HR, hazard ratio

*National Bureau of Statistics of China. China Population and Employment Statistics Yearbook 2020-2023.

**Table S14. Annual transition probabilities and HCC/LT rates**

| From/To | Steatosis | ASH | Fibrosis | Cirrhosis | DC | HCC | LT | Death |
| --- | --- | --- | --- | --- | --- | --- | --- | --- |
| Healthy | 0.012 | N/A | N/A | N/A | N/A | N/A | N/A | 0.005 |
| Steatosis | N/A | 0.03 | 0.08 | N/A | N/A | N/A | N/A | 0.06 |
| ASH | N/A | N/A | 0.10 | 0.08 | N/A | N/A | N/A | 0.11 |
| Fibrosis | N/A | N/A | N/A | 0.08 | 0.04 | 0.01-0.04 | N/A | 0.08 |
| Cirrhosis | N/A | N/A | N/A | N/A | 0.07-0.08 (1^st^ year) | 0.01-0.04 | 0.001-0.005 | 0.15-0.20 (1^st^ year) |
| DC | N/A | N/A | N/A | N/A | N/A | 0.005-0.02 | 0.001-0.005 | 0.10-0.15 |
| HCC | N/A | N/A | N/A | N/A | N/A | N/A | 0.10 | 0.40-0.60 (1^st^ year) |
| LT | N/A | N/A | N/A | N/A | N/A | N/A | N/A | 0.05-0.10 (1^st^ year) |

ALD, alcohol-associated liver disease; ASH, alcoholic steatohepatitis; DC, decompensated cirrhosis; HCC, hepatocellular carcinoma; LT, liver transplantation

**Table S15. Subgroup prevalence of ALD**

| **Year** | **Male (%)** | **Female (%)** | **Urban (%)** | **Rural (%)** |
| --- | --- | --- | --- | --- |
| 2020 | 7.1 | 2.5 | 5.2 | 4.4 |
| 2025 | 8.3 | 2.8 | 6.0 | 5.0 |
| 2030 | 9.5 | 3.0 | 6.8 | 5.6 |
| 2035 | 10.4 | 3.3 | 7.4 | 6.2 |
| 2040 | 11.2 | 3.6 | 8.0 | 6.8 |
| 2045 | 11.8 | 4.2 | 8.6 | 7.3 |
| 2050 | 12.3 | 4.7 | 9.2 | 7.8 |

ALD, alcohol-associated liver disease
